# Depressive rumination is correlated with brain responses during self-related processing

**DOI:** 10.1101/2021.03.18.21253930

**Authors:** Tzu-Yu Hsu, Tzu-Ling Liu, Paul Z. Cheng, Hsin-Chien Lee, Timothy J. Lane, Niall W. Duncan

**Affiliations:** Graduate Institute of Mind, Brain and Consciousness, Taipei Medical University, Taipei, Taiwan; Brain and Consciousness Research Centre, TMU Shuang-Ho Hospital, New Taipei City, Taiwan; Institute of Cognitive Neuroscience, National Central University, Zhongda, Taiwan; Department of Psychiatry, School of Medicine, College of Medicine, Taipei Medical University, Taipei, Taiwan; Department of Psychiatry, Taipei Medical University Hospital, Taipei, Taiwan

**Keywords:** rumination, self, EEG, major depressive disorder, negative thought

## Abstract

**Background:** Rumination, a tendency to focus on negative self-related thoughts, is a central symptom of depression. Studying the self-related aspect of such symptoms is challenging due to the need to distinguish self effects *per se* from the emotional content of task stimuli. This study employs an emotionally neutral self-related paradigm to investigate possible altered self processing in depression and its link to rumination.

**Methods:** People with unipolar depression (MDD; n = 25) and controls (n = 25) underwent task-based EEG recording. Late event-related potentials were studied along with low frequency oscillatory power. EEG metrics were compared between groups and correlated with depressive symptoms and reported rumination.

**Results:** The MDD group displayed a difference in late potentials across fronto-central electrodes between self-related and non-self-related conditions. No such difference was seen in controls. The magnitude of this difference was positively related with depressive symptoms and reported rumination. MDD also had elevated theta oscillation power at central electrodes in self-related conditions, which was not seen in controls.

**Conclusions:** Rumination appears linked to altered self-related processing in depression, independently of stimuli-related emotional confounds. This connection between self-related processing and depression may point to self-disorder being a core component of the condition.

## Introduction

Rumination involves repetitive thoughts about the feelings and problems that one has (Smith and Alloy, 2009). It is a common symptom in major depressive disorder (MDD), where the thoughts tend to involve a focus on negative emotions and life-problems. This distinguishes depressive rumination from constructive repetitive thought, which can be of benefit to the individual (Watkins, 2008). The tendency towards negative rumination in MDD is correlated with the severity and duration of depressive episodes (see Nolen-Hoeksema et al., 2008 for a review), as well as with suicide risk (Holdaway et al., 2018), and with the likelihood of relapse in those recovering from the condition (Michalak et al., 2011; Spinhoven et al., 2018). Rumination also appears to be a risk factor in healthy individuals for developing MDD (Spasojević and Alloy, 2001) and to predict its onset (Abela and Hankin, 2011). Investigating the root neural changes leading to maladaptive rumination is thus an important step towards understanding MDD and individual differences in vulnerability to that condition.

A fundamental feature of rumination is that the thoughts involved are self-related. Based on this, a relationship between the mechanisms of rumination and the mechanisms of self-related thought has been proposed (Nejad et al., 2013). Such a supposition is supported by an overlap in the brain regions involved in processing each. More specifically, self-related thought has been linked to brain regions along the cortical midline (cortical midline structures; CMS) such as the medial prefrontal and posterior cingulate cortices (Qin and Northoff, 2011). Activity properties in these same regions have then been further linked to individual differences in general rumination and are modulated by a rumination task (Berman et al., 2011; Zamoscik et al., 2014; Zhu et al., 2012). These cortical midline structures have also been linked to depressive symptoms and negative self-focused thought (Kaiser et al., 2016; Philippi et al., 2018). Finally, activity within CMS during self-referential thinking appears to differ between MDD patients and controls (Grimm et al., 2009; Lemogne et al., 2010; Yoshimura et al., 2010). Together, this evidence suggests that investigating changes to self-related processing may provide important insights into ruminative symptoms in MDD.

Investigating self-related processing in MDD presents a methodological challenge as many of the tasks used involve stimuli that have emotional valence in addition to being self-related. For example, a commonly used task asks participants to relate particular positive or negative personal traits to themselves or to others (e.g., Lemogne et al., 2010; Yoshimura et al., 2010). This task induces a late positive potential (LPP) over fronto-central regions that differs between patients with depression and controls (Auerbach et al., 2016, 2015; Shestyuk and Deldin, 2010). The emotional valence of the stimuli used in such tasks may, however, interact with the negative emotional bias found in depression (Disner et al., 2011). This means that people with depression tend to focus more on negative self-related stimuli and interpret stimuli more negatively in relation to themselves (Gotlib et al., 2004; Mennen et al., 2019; Miskowiak et al., 2018). With this potential interaction between the self-related stimuli employed and an inherent negative bias, it becomes challenging to parse the contribution to brain activity measures of self-related processing *per se* from the influence of altered emotional processing.

Tasks that use emotionally neutral stimuli may circumvent this issue and allow more direct study of self-related processing in MDD. One such task was developed by Johnson and colleagues (Johnson et al., 2005) in which sets of three coloured patches are used as the stimulus. Participants are asked to judge which pair of colour patches they prefer in one task condition while in the other they are asked which pair is most similar in terms of their relative positions in colour space. These two conditions respectively represent self-related (subjective: “which do *I* prefer”) and non-self-related (objective: “which is most like the other”) judgements. An increase in central frontal activity is observed in the self-related condition, in line with the described relationship between CMS and self-related processing (Johnson et al., 2005; Nakao et al., 2013a, 2013b). These brain responses are likely to be independent of task-induced emotional biases and so provide a tool to investigate the relationship between self-related processing and rumination without also probing negative bias.

The current study took advantage of this colour judgement task, in conjunction with electroencephalogram (EEG) recordings, to investigate the association between self-related processes and rumination in a group of MDD patients, along with age and sex matched healthy controls. Ruminative behaviours were assessed with the Ruminative Responses Scale (RRS; Treynor et al., 2003). An ERP approach was used to analyse the EEG data, focussing on the LPPs identified in prior work. It was predicted that self and non-self ERPs would differ between patients and controls at central-frontal electrodes and that these differences would correlate with RRS scores. This approach advances prior work linking self-related processes and rumination into the temporal domain in a manner that may be less biased by differences in emotional processing.

## Methods

### Participants

Twenty-five patients diagnosed with MDD (20 females; 38.6 ± 13.5 years old) were recruited at the Shuang-Ho Hospital Department of Psychiatry. Diagnosis was made according to the Diagnostic and Statistical Manual of Mental Disorders, Fourth Edition. A MINI structured interview (Sheehan et al., 1998) was used for diagnosis confirmation, detection of suicide risk, and to exclude patients with comorbid mental or substance use disorders. Patients with poor visual acuity or a history of neurological issues (e.g. stroke, seizure, traumatic brain injury, post-brain surgery) were also excluded. The average duration of the most recent depressive episode was 4.1 ± 3.15 months. All patients but one were on medication at the time of testing (see Table S1 for details). Twenty-five healthy control participants (21 females, 38.2 ± 14.2 years old) without any current or history of neurological or psychiatric disorders, and who were not using any psychotropic medication, were also recruited from the community. Controls and patients were matched for age and sex, apart from in one case where the sex was different. The study was approved by the TMU Institutional Review Board (N201603080). Written consent was obtained from all participants prior to their participation in this study. All participants were informed about the purpose of the study and procedures before giving this.

### Questionnaires

Ruminative behaviours were assessed in both patients and controls with the 22 item Ruminative Response Scale (RRS; Treynor et al., 2003). This contains three subscales - reflective, brooding, and depressive rumination. The reflective and brooding subscales were focused on here as these separate behaviours into adaptive reflective thoughts and maladaptive brooding thoughts (Schoofs et al., 2010; Treynor et al., 2003). Total scores plus each of these subscores were calculated for each participant. The 21 item Beck Depression Inventory (BDI-II; Beck et al. 1996) was administered to both patients and controls to evaluate the presence of general depressive symptoms across both groups. Both questionnaires were presented in Traditional Chinese (Huang et al., 2015; Lu, 2002). Patients had significantly higher BDI and RRS scores than controls (see Table 1).

**Table 1.** Mean (SD) Beck Depression Index (BDI) and Rumination Response Scale (RRS) scores for MDD and control groups.

| Scale | Control | Patient | Comparison |
| --- | --- | --- | --- |
| BDI | 7.8 (7.7) | 28.7 (14.4) | $t = -6.7, p < .001$ |
| Rumination | 40.4 (12.5) | 62.8 (14) | $t = -6.1, p < .001$ |

### Colour judgement task

The general configuration of the task stimuli is illustrated in Figure 1. Each trial began with a 1500∼2000 ms fixation cross (subtending 0.5° × 0.5°). A stimulus array consisting of three non-overlapping, differently coloured squares was then presented for 2000 ms. One square was located at the middle top, with the remaining two located on the left and right bottom. The top square was the target to which the other two were compared in each trial. Each coloured square subtended 1.5° × 1.5° visual angle.

**Figure 1:**
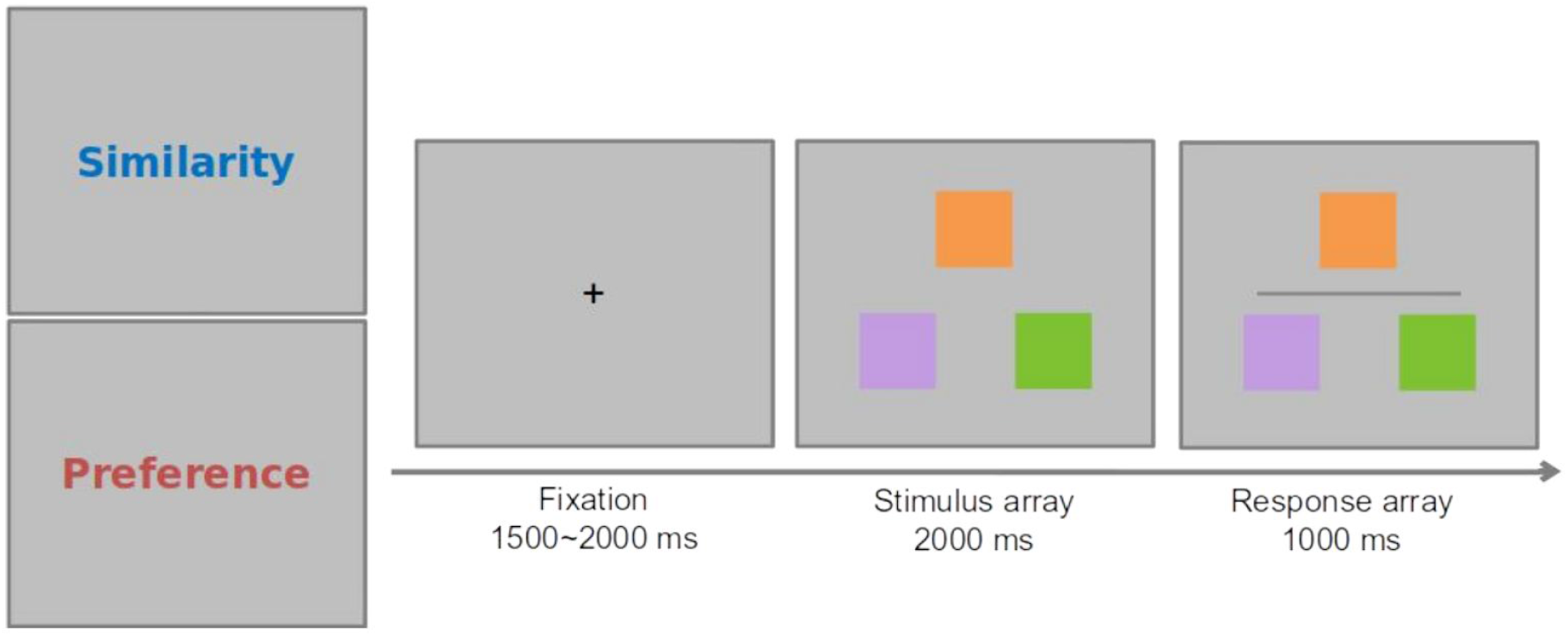
Participants were instructed at the start of each block to make either a similarity or preference judgment. Each trial started with a 1500 to 2000 ms fixation period, followed by the stimulus and response arrays. The stimulus array consisted of three colour swatches upon which participants made their judgement. A dark grey horizontal line then appeared to indicate the start of the response period, during which participants pressed either the left or right button to indicate their decision.

Three colours were selected for each trial from a set of 12 taken from the CIELAB colour space at 30° intervals around the a*(red-green)-b*(yellow-blue) plane colour wheel. Trial difficulty was determined by the objective similarity in hue between the target and the two lower squares. In low difficulty trials, one choice colour would be 60° from the target colour and the other 150°. In high difficulty trials, the relative difference in distance would be less (90° and 120°), making it harder to discriminate the degree of similarity to the target. The luminance of the three squares was equalised by manipulating the red, green, and blue (RGB) components of the colours.

The task consisted of two different conditions. During the colour similarity judgement condition, participants were required to judge which of the lower squares was most similar in hue to the target and then press the corresponding left or right button on the mouse (Figure 1). For the preference judgement condition, participants were instructed to choose the colour pairing that they preferred between the target and either of the lower squares. The same low and high difficulty sets of colours were used in the preference trials as in the similarity ones. Following the viewing period, participants were required to respond within 1000 ms. This response period was indicated by a dark gray horizontal line appearing on the screen.

Participants completed 20 practice trials before performing 4 blocks of 48 trials. The preference judgment and similarity judgement conditions were block design. Participants were instructed which condition they were going to perform before each block. Each block was initiated by the participant, allowing for breaks between blocks to reduce fatigue. Participants were instructed to maintain visual fixation at the center of the screen while avoiding blinks and eye movements during each trial. The sequence of conditions was counterbalanced across participants.

The experiment was programmed in Psychtoolbox 3 (Brainard, 1997) running on MATLAB R2015a (MathWorks). Stimuli were displayed on a 24-inch CRT monitor with a vertical refresh rate of 60 Hz. Participants viewed the stimuli at a distance 57 cm, with a chin-rest used to stabilize their head position.

### EEG recording and preprocessing

EEG was recorded with Ag/AgCl electrodes mounted in an elastic cap (easycap, Brain Products GmbH) using a 30-electrode arrangement following the International 10-20 System, including mono-polar electrodes (FP1/2, F7/3/Z/4/8, FC5/3/Z/4/6, T7/8, C3/Z/4, CP5/1/Z/2/6, P7/3/Z/4/8, O1/Z/2). An additional 6 channels were attached for measuring eye movement and as references. Vertical eye movement was recorded with two electrodes on the supraorbital and infraorbital ridges of the left eye and horizontal with two electrodes on the outer canthi of the right and left eyes. The remaining two electrodes attached to mastoid sites served as the reference (A1 and A2). Electrode impedances were kept below 10 kΩ for all electrodes. Online EEG was recorded with BrainAmp with a bandpass filter of 0.05-1000 Hz. Data was recorded with BrainVision Recorder 1.2 software, with a sampling rate of 1000 Hz.

For off-line analysis, EEG preprocessing was conducted using EEGLAB (Delorme and Makeig, 2004, RRID:SCR_007292) and ERPLAB (Lopez-Calderon and Luck, 2014, RRID:SCR_009574) MATLAB toolboxes. Continuous data were first low-pass filtered at a cut-off frequency of 30 Hz. After manually excluding noisy periods, independent component analysis (ICA) was conducted on the continuous waveforms to identify signals caused by eye movement, blinks, and changes in muscle tone. The contribution of these task irrelevant signals to the data was removed using the ICA in EEGLAB. Continuous data were then segmented into 2200 ms epochs, including 200 ms pre- and 2000 ms post-stimulus intervals. Epochs were baseline-corrected with the pre-stimulus interval. Finally, epochs that contained signals exceeding ±60 mv in any channel were rejected and the remaining trials averaged into “similarity-low,” “similarity-high,” “preference-low,” and “preference-high” categories (Table 2).

**Table 2.** Mean (SD) behavioural responses in each task condition for control and MDD groups.

|  | Similarity |  | Preference |
| --- | --- | --- | --- |
|  | Low | High | --- |
| Accuracy |  |  |  |
| Control | 0.89 (0.07) | 0.64 (0.11) | --- |
| MDD | 0.84 (0.15) | 0.59 (0.16) | --- |
| Reaction times (ms) |  |  |  |
| Control | 627 (107) | 647 (100) | 637 (102) |
| MDD | 646 (104) | 664 (97) | 655 (99) |

### Behavioural data analysis

Performance during similarity trials in terms of reaction times and accuracy was compared using a 2 (difficulty) x 2 (group) mixed ANOVA. Preference trials had no objective criteria and so their accuracy could not be analysed. As such, reaction times for preference trials were compared between groups through an independent sample t-test. Finally, any interaction between condition and group for reaction times was tested through a 2 (condition) x 2 (group) mixed ANOVA. Behavioural data for one control participant was not available.

### Analysis of task-induced response differences

Event-related potentials (ERP) induced by the task were analysed to identify any overall differences in brain responses between patients and controls. The 900-2000ms time-period was tested, covering sustained self-related components (Auerbach et al., 2016, 2015; Herbert et al., 2011). Responses were averaged within this period at twelve electrodes located along the midline and in the left and right hemispheres at the frontal (Fz/F3/4), central (Cz/C3/C4), parietal (Pz/P3/P4) and occipital (Oz/O1/O2) scalp. These were then compared using a mixed ANOVA that included a between-subject factor of group (control, MDD) and within-subject factors of condition (similarity, preference), difficulty (low, high), and anterior-posterior location (frontal, central, parietal, occipital). Post-hoc tests were used to identify which anterior-posterior locations showed a difference between conditions in each group. P-values were corrected for sphericity according to Greenhouse-Geisser adjustment. Holm–Bonferroni adjustment was applied to control the family-wise error rate. Statistical analyses were conducted using JASP (JASP Team, 2020, RRID:SCR_015823).

Groups of electrodes where group*condition interactions were observed were analysed further to establish more precisely the times and locations at which differences between conditions occur. Waveforms from individual electrodes within the selected electrode groups were compared at each timepoint between conditions through paired t-tests. Threshold-free cluster-based thresholding was used to identify periods of time across multiple electrodes where conditions differed (Mensen and Khatami, 2013).

### Self vs non-self response differences and depressive rumination

Having established the specific time periods and electrodes at which activity differed between similarity and preference conditions, the relationship between activity in these periods and depressive rumination was then investigated. The timepoints at specific electrodes where a difference between conditions was seen in the MDD group was used as a mask to extract average activity from both that group and controls. These averaged activity levels were then correlated with BDI scores in each group separately (Spearman’s correlation). The difference between correlation strength was also tested through linear regression. A similar analysis was then conducted using RRS scores.

### Self vs non-self related processes and midline oscillatory power

In a final, exploratory step, induced response power at the fronto-central midline electrodes with LPP differences (Fz and Cz) was calculated, focusing on the alpha and theta bands. These frequency ranges were selected given their suggested connections to self-related processing (Knyazev, 2013). Power spectral density was calculated for the period from stimulus onset to 2000 ms. Oscillatory power was separated from the 1/f aperiodic component of the signal using the Fooof Python toolbox (Donoghue et al., 2020). Theta power was averaged within the 4-8 Hz range and alpha from 8-13 Hz. Mixed-effect linear regression was then used to examine an oscillatory power fixed-effect group by condition interaction (with subject as a random effect). Holm–Bonferroni adjustment was used to correct *p*-values for the two electrodes and two frequency bands analysed.

## Results

### Behavioural results

In the similarity task, participants were more accurate during the low difficulty than in the high (86.5% vs 60.5%; *F*(1,47) = 186.04, *p* < 0.001). Accuracy did not differ between the groups (*F*(1,47) = 2.54, *p* = 0.12), nor was there an interaction between group and difficulty (*F*(1,47) = 0.023, *p* = 0.87). Reaction times in the similarity task were faster in the low difficulty condition than in the high (636 vs 655 ms; *F*(1,47) = 16.1, *p* < 0.001). Reaction times in the similarity task were faster in the easy than in the difficult condition (636 vs 655 ms; *F*(1,47) = 16.1, *p* < 0.001). No effect of group on reaction time was found (*F*(1,47) = 0.37, *p* = 0.55), nor was there an interaction between difficulty and group (*F*(1,47) = 0.038, *p* = 0.84). Reaction times in the preference task did not differ between groups (*t*(47) = 0.73, *p* = 0.46). No interaction between task and group was found for reaction times (*F*(1,47) = 0.037, *p* = 0.84). All response accuracies and reaction times are given in Table 2.

### Task-induced response differences

Task-induced ERP amplitudes during the 900-2000 ms period are summarized in Figure 2. No difficulty*condition*group*location interaction was observed (*F*(1.51, 72.25) = 0.76, *p* = 0.44). As such, the difficulty factor was collapsed for all subsequent analyses. An interaction between condition, group, and location was then observed (*F*(1.5, 71.77) = 4.20, *p* = 0.03). Follow-up simple main effect analyses highlighted differences at the frontal and central locations. More specifically, the MDD group displayed a more negative waveform during the similarity condition, compared to preference, at the frontal (*p* < 0.001) and central areas (*p* = 0.001) but not parietal (*p* = 0.44) and occipital (*p* = 0.06). In contrast, the control group showed no amplitude differences between these conditions at any location (frontal: *p* = 0.12; central: *p* = 0.17; parietal: *p* = 0.11; occipital: *p* = 0.89).

**Figure 2:**
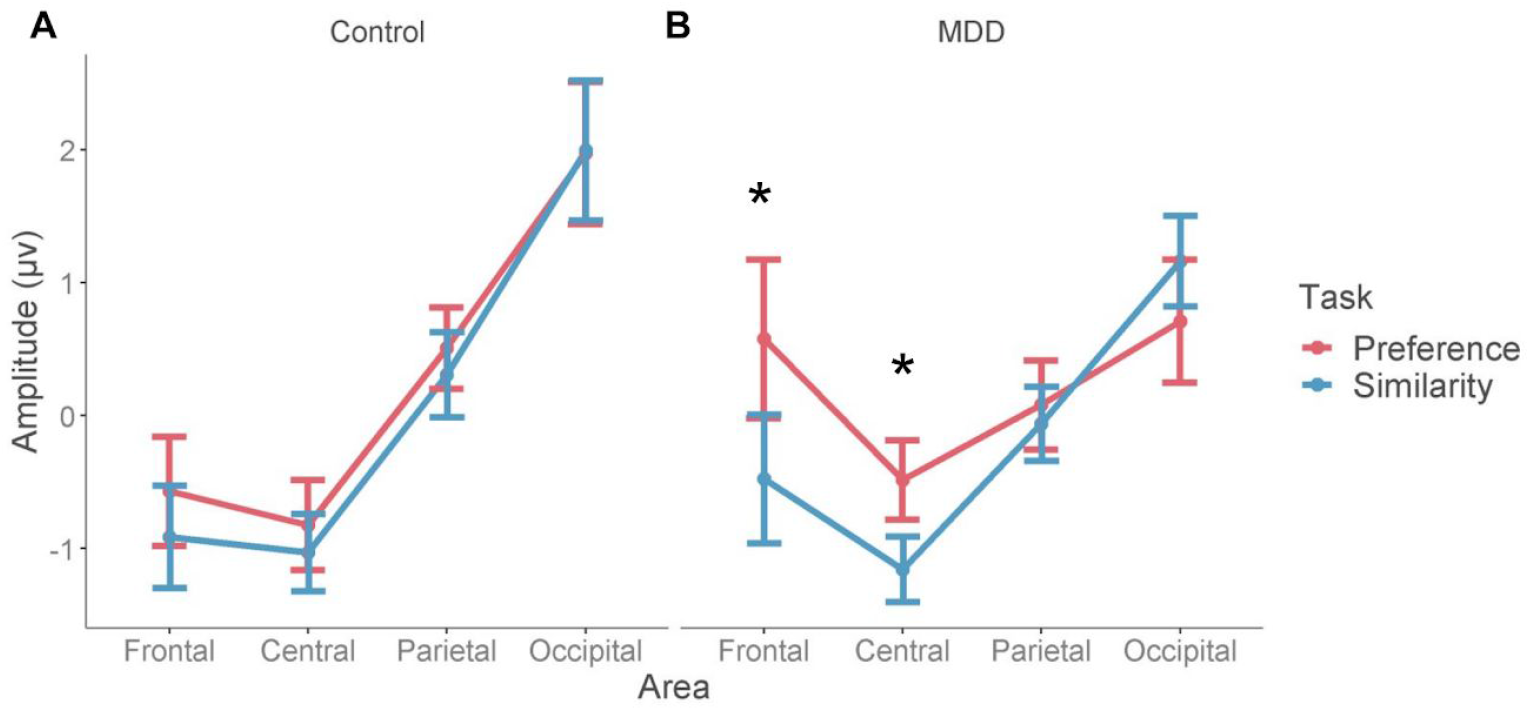
Mean ERP amplitudes at different scalp areas from 900 to 2000 ms after the stimulus array onset, shown separately for the (A) control and (B) MDD groups. Responses during the similarity condition are shown in blue and those during the preference condition in red. Differences between conditions were seen at frontal and central areas in the MDD but not control group. Error bars represent the standard error of the mean. * indicates p < 0.05.

Investigating the frontal and central locations in more detail, comparisons between conditions at each time point and each relevant electrode were conducted. Significant differences between preference and similarity waveforms were observed for the MDD group that start at the frontal electrodes around 815 ms and then propagate to the central area (Figure 3A). During this period, measured activity became less negative over time for the preference condition than similarity. No differences between conditions were seen for the control group (Figure 3B).

**Figure 3:**
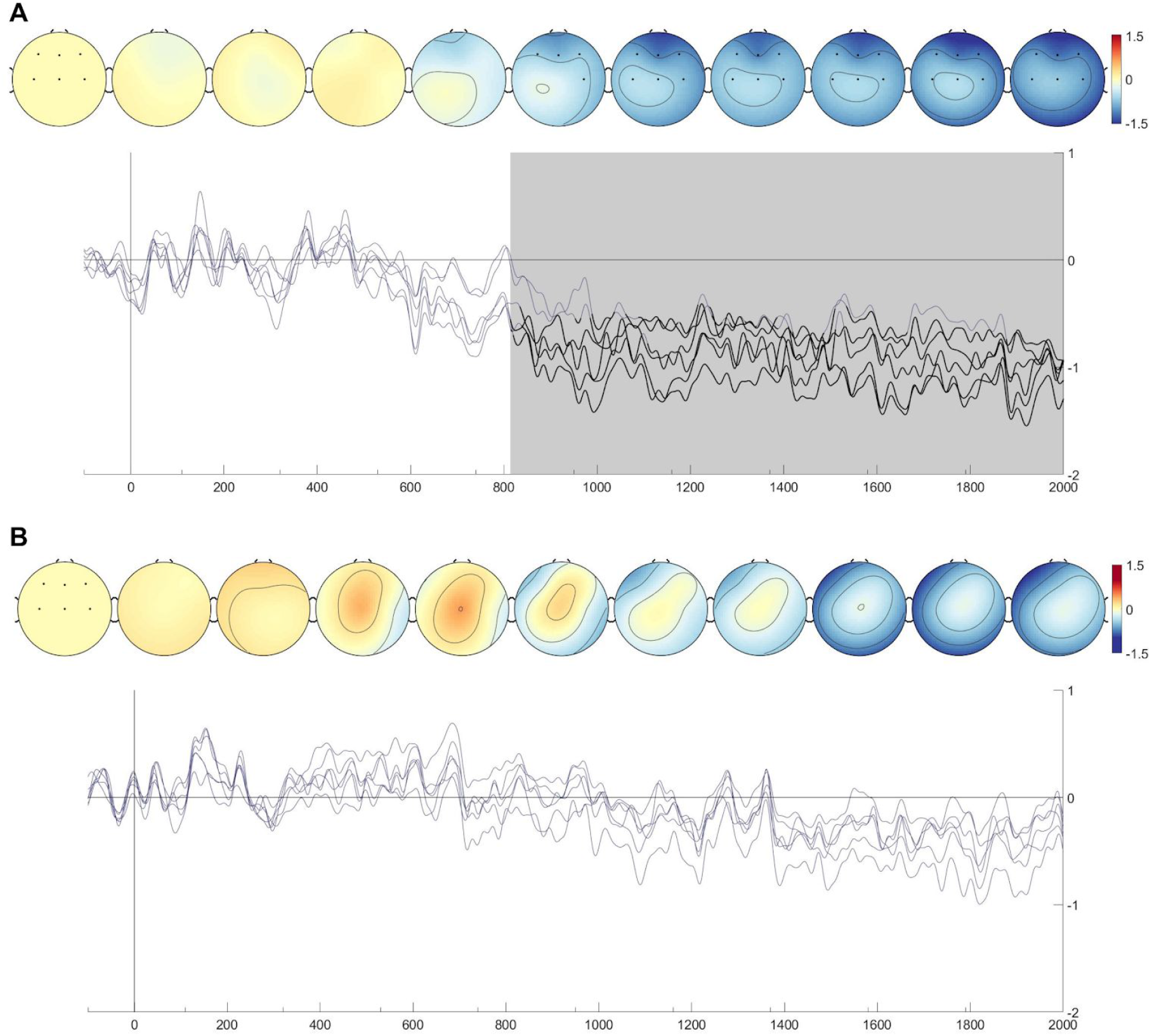
Similarity-Preference differences for (A) MDD and (B) control groups. Electrodes for which comparisons were done are indicated in black on the topographical maps. These maps show the Similarity-Preference difference over time. Waveforms represent Similarity-Preference differences for each selected electrode. Significant differences per electrode are shown in black. Periods where at least one difference is found are shaded in grey. Visualizations were created using the EEGVIS toolbox (Ehinger, 2018).

### Self vs non-self response differences and depressive rumination

As shown in Figure 4A, the difference in amplitude between the similarity and preference conditions was positively correlated with BDI scores in the MDD group (*r*(24) = 0.61, *p* = 0.001) but not in the control group (*r*(24) = -0.36, *p* = 0.075; Figure 4A). The slopes of these correlations differed from each other (*t*(46) = 3.01, *p* = 0.004). A similar correlation between RRS scores and amplitude differences was also seen in the MDD group (*r* = 0.48, *p* = 0.016) but not controls (*r*(24) = -0.13, *p* = 0.54; Figure 4B). These correlations differed from each other (*t*(46) = 2.43, *p* = 0.019).

**Figure 4:**
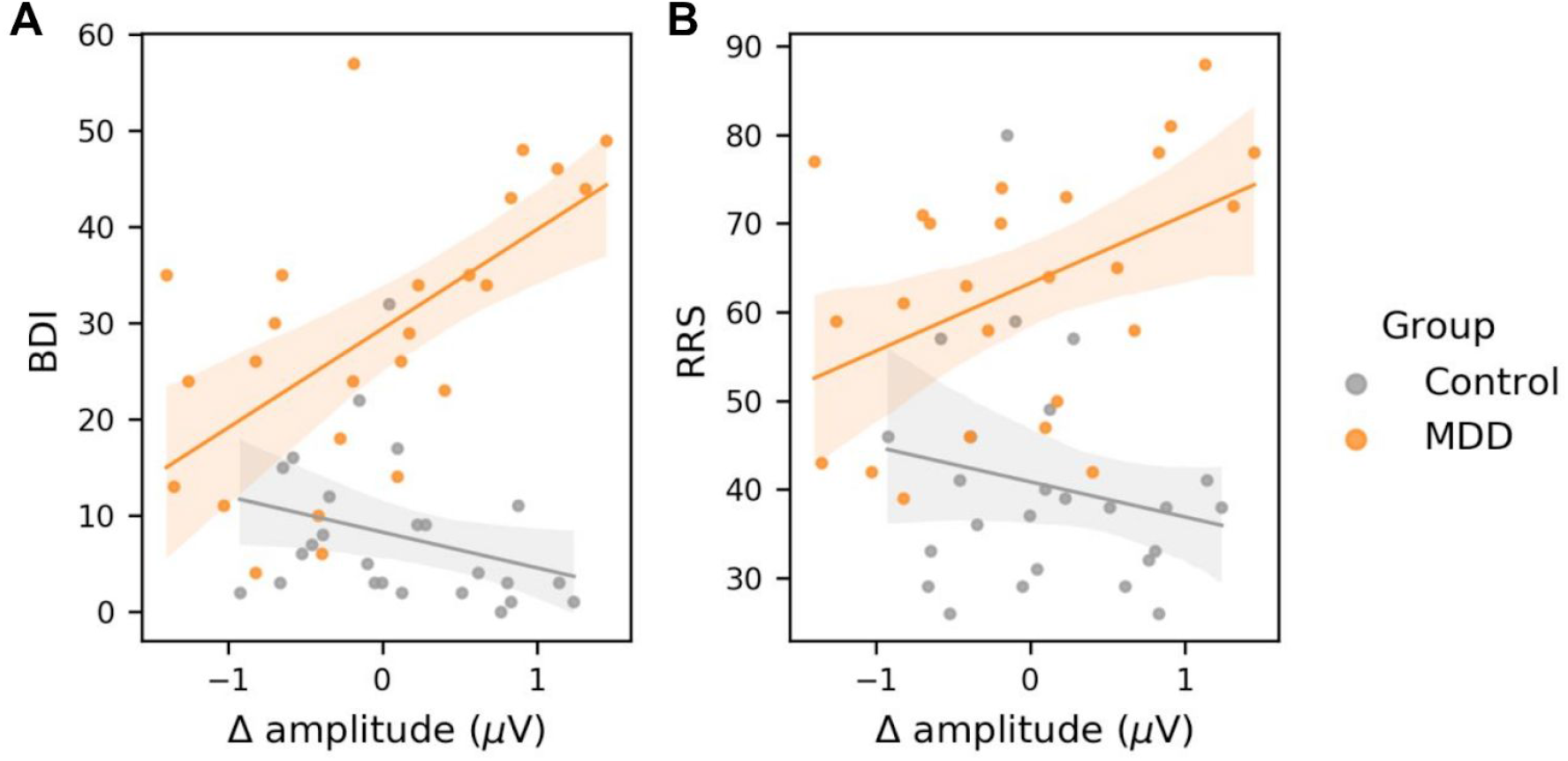
Differences in the amplitude of sustained responses between preference and similarity conditions were correlated with (A) BDI and (B) RRS scores. Data from the MDD group are shown in orange and those from the control group in grey. Significant relationships between amplitude differences and scores were observed for MDD but not control participants.

### Self vs non-self related processes and midline oscillatory power

Analysing the interaction between task condition and group, no effects were observed at Fz for oscillatory power within either the theta (*β* = 0.033, *z* = 0.4, *p* = 1; Figure S1) or alpha bands (*β* = -0.023, *z* = -0.61, *p* = 1; Figure S2). In contrast, an interaction between condition and group was observed within the theta band at Cz (*β* = -0.12, *z* = 2.92, *p* = 0.016; Figure 5). No such effect was seen for alpha power at this electrode (*β* = -0.002, *z* = -0.054, *p* = 1; Figure S3). Post-hoc t-tests (Holm-Bonferonni adjusted) showed that MDD patients had higher theta power in the preference compared to similarity condition (*t*(24) = -2.65, *p* = 0.038). Theta power did not differ between conditions for control participants (*t*(24) = 1.08, *p* = 0.6).

**Figure 5:**
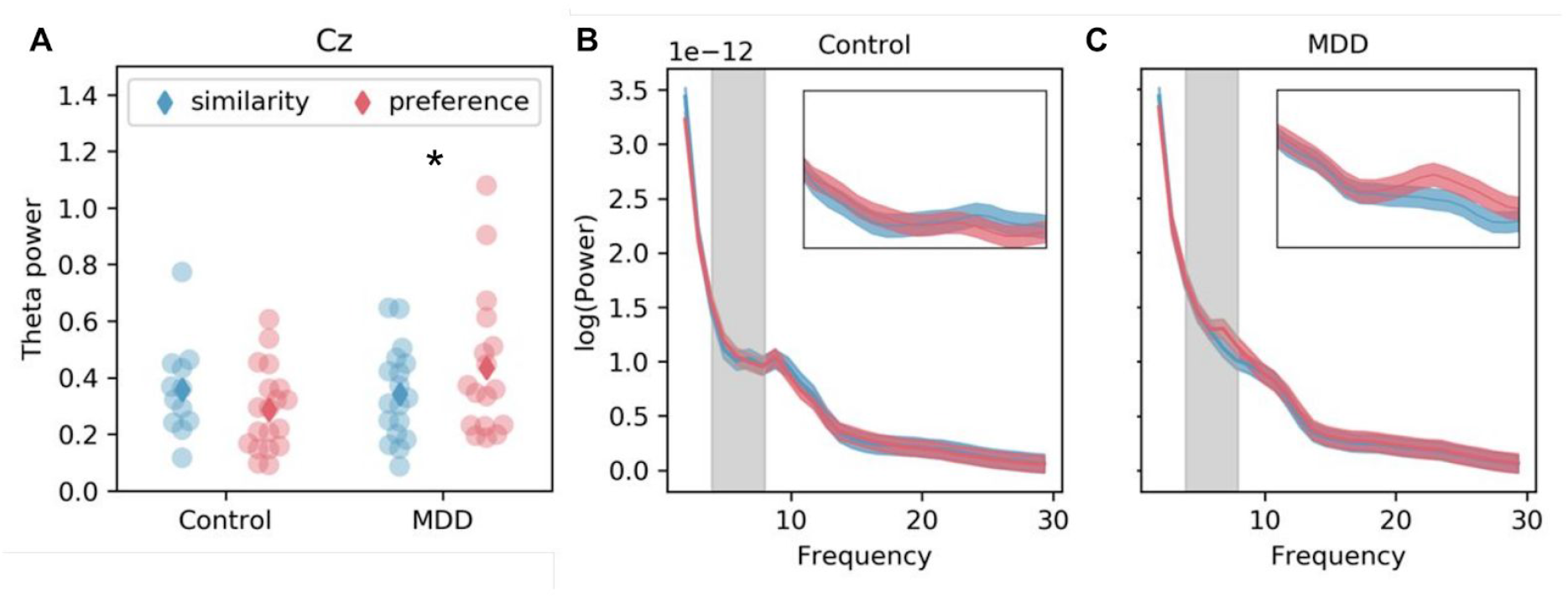
(A) Theta band oscillatory power at Cz as a function of condition in MDD and control groups. Dots represent values from individuals in the similarity (blue) and preference (red) conditions. Group means are indicated by diamonds. Power spectra from which values are obtained are shown for control (B) and MDD (C) groups. Insets show a close-up of the theta range indicated by grey shading in the main spectra. * indicates p < 0.05.

## Discussion

The relationship between self-related activity and depressive rumination was investigated through an emotionally neutral task in patients with MDD and healthy controls. Differences between groups in a late ERP component were seen at frontal and central electrodes. MDD patients displayed less negative activity than controls during the subjective preference condition than the objective similarity condition. This amplitude difference between the similarity and preference conditions was positively correlated with BDI and RRS scores in the MDD group. Moreover, it was shown that an increase in central theta power occurs during the preference condition in participants with MDD.

In terms of behavioural performance, no difference in accuracy and reaction times for the similarity or preference conditions was observed between groups. Reaction times were faster in low difficulty trials than high difficulty ones. This sensitivity to trial difficulty suggests that participants engaged with the task appropriately. This was equally true for both patients with MDD and controls, as demonstrated by the lack of interaction between difficulty and group. Taken together, these behavioural measures point to the colour similarity and preference task being completed equally well by both patients and controls.

Sustained late ERPs at frontal and central electrodes were observed in the current work. Previous ERP studies of self-related processing have identified similar LPPs. Those experiments employed a self-reference task in which participants attribute positive or negative words to themselves (Kuiper and Derry, 1982). In such tasks, patients with depression tend to link negative words to themselves whilst controls select more positive ones (Lemogne et al., 2010). At the level of LPPs, patients display greater amplitude responses for negative than positive words over fronto-central regions. This is in contrast to controls, where positive words induce greater evoked responses (Auerbach et al., 2016, 2015; Shestyuk and Deldin, 2010). In patients, the magnitude of the difference between LPPs induced by positive and negative words appears to increase with symptom severity (Benau et al., 2019), suggesting that people with depression may selectively elaborate upon negative emotional self-related information. Notably, and in contrast to these prior studies, the task used here does not involve an emotional component. The presence of these sustained late ERPs in the context of this task thus suggests that LPP-like effects in general may be driven, in part at least, by self-related processes that are independent of emotional content.

In this study, a sustained difference in LPP-like activity between self-related and non-self-related stimuli in patients was seen until trial-end. This was not seen in controls, where no such sustained difference was observed during this period. Notably, the difference in this activity between conditions was positively correlated with reported ruminative behaviours in the MDD group. MDD patients’ sustained LPP-like reactivity in an emotionally-neutral context may point to people with depression being inclined towards engaging in continual thought whose nature is fundamentally self-related. This would be distinct from thought that is primarily emotional in nature and which therefore also has a self-related aspect as a secondary feature. This is in line with prior work linking rumination in depression to activity within regions, such as the mPFC and PCC (Hamilton et al., 2015; Sheline et al., 2009), that are activated by self-related processes independent of emotion (Qin et al., 2020). A connection between mPFC and PCC activity and this LPP-like activity is further supported by these regions being more activity during the preference condition in prior work that has employed the task in fMRI (Johnson et al., 2005). These potential associations may merit further research with tools that can better resolve temporal and spatial properties, such as MEG or high density EEG.

An analysis of oscillatory power at midline fronto-central electrodes revealed elevated Cz theta power during the preference condition in the MDD group specifically. A range of prior work has found associations between theta band activity and self-related processing (Knyazev, 2013). This work also provides evidence for a connection between theta activity and CMS. That only patients demonstrate an increase in theta power during the self-related preference condition suggests that there may be a greater sensitivity in depression to the subjectivity of experiences, generating a response that may only be seen in non-depressed individuals with explicit self cueing (Mu and Han, 2013). This sensitivity may then lead to an increased likelihood to engage in self-related thought in a manner leading to ruminative symptoms. Such a suggestion remains speculative, however, and requires further investigation.

A number of limitations should be noted. The first of these was the medication status of participants, where all but one were undergoing pharmacological treatment. One previous study has found an influence of SSRIs on self-reflective activity but this highlighted subcortical regions and did not reveal any effect upon cortical regions such as the mPFC or PCC (Young et al., 2020). This suggests that the effects seen here are not driven by drug effects but further research is required to confirm this. Secondly, some prior self-related tasks, such as the self-referential one noted above, have overlapped judgements and responses, potentially leading to motor induced contamination. This was not the case in the present study as two seconds clearly separated responses from the judgement period. This means that differences between groups are unlikely to be due to motor preparation or execution effects. Finally, our participant group was primarily female. There is some evidence for sex-differences in the relationship between self-related thought and depressive symptoms, where there may be less of an association in males (Fast and Funder, 2010). The current results therefore require replication in a balanced participant sample.

Taken together, the current findings suggest that rumination is related to altered self-related processing in depression and that this can be shown independently of potential confounds arising from task stimuli with explicit emotional content. The difference in neural responses induced by self-related stimuli in MDD patients lends weight to a conception of MDD that includes self-disorder as a fundamental component. Future research may wish to focus on treatment options that engage more directly with the self, including pharmacological interventions such as psilocybin that produce self-dissolution effects (Mason et al., 2020).

## Data Availability

Participants did not give consent to their data being shared openly and so the data included in this analysis are only available from the authors upon reasonable request.

## Acknowledgements

The authors would like to thank all participants for their time and effort. They are grateful also to Hsin-Yi Wang and Ching Lin for help with patient recruitment and data collection. This work was supported by funding from the Taiwan Ministry of Science and Technology to TJL (107-2632-H-038-001-MY3), TYH (109-2410-H-038-010), and NWD (108-2410-H-038-008-MY2). This work was also supported by the Taiwan Ministry of Education Higher Education Sprout Project.

## Competing interests

The authors declare no conflicts of interest.

## Author contributions

TYH, TLL and NWD analysed the data; PZC and HCL collected data; TYH, NWD and TJL designed the study; TYH, TLL and NWD wrote the manuscript; all authors revised and approved the manuscript.

---

**Supplementary Table 1:**
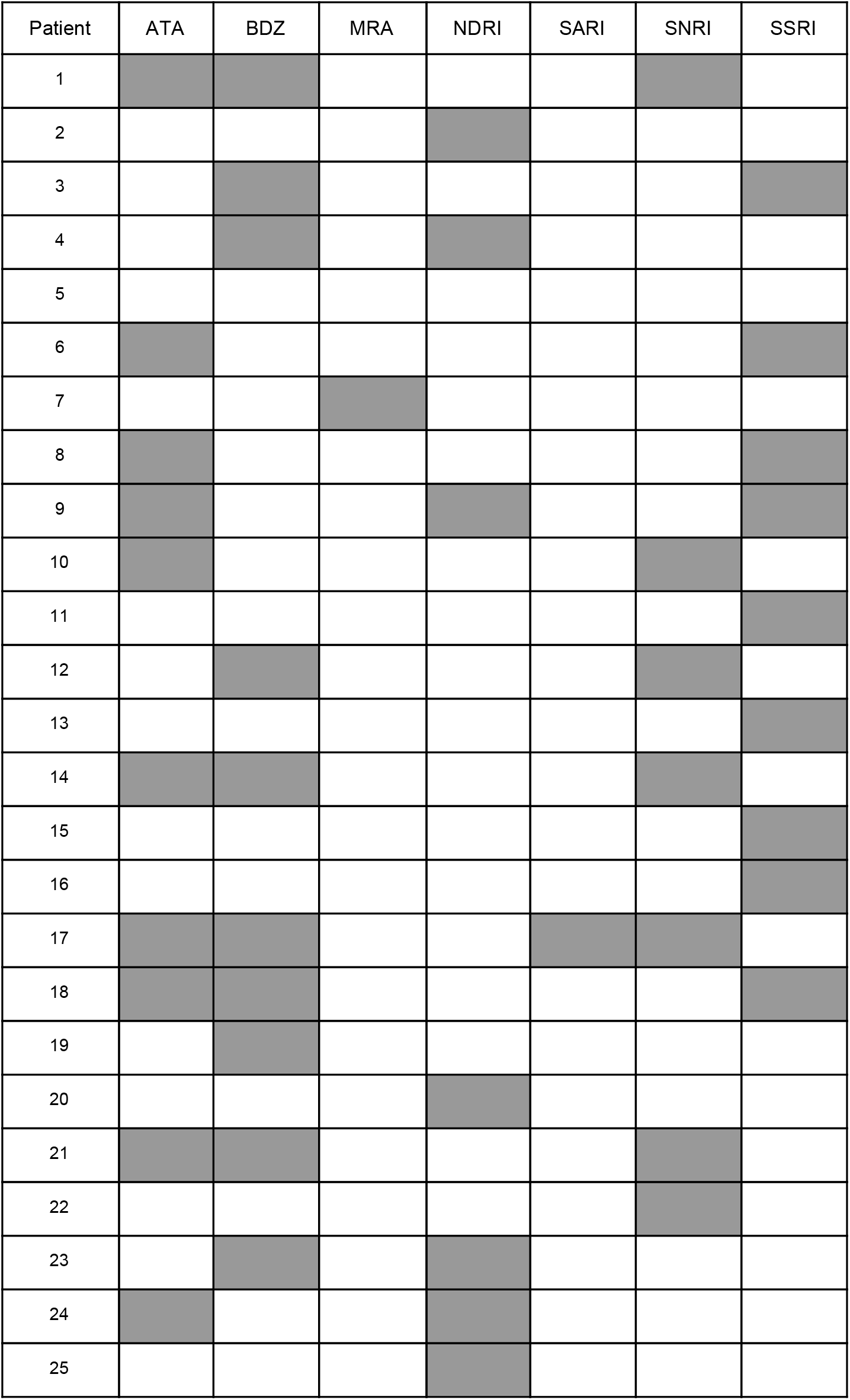
Medications for MDD patients. Shadowed boxes indicate that a patient was taking that medication at the time of the experiment. ATA = Atypical antipsychotic; BDZ = Benzodiazepine; MRA = Melatonin receptor agonist; NDRI = Norepinephrine-dopamine reuptake inhibitor; SARI = Serotonin antagonist and reuptake inhibitor; SNRI = Serotonin-norepinephrine reuptake inhibitor; SSRI = Selective serotonin reuptake inhibitor.

**Supplementary figure 1:**
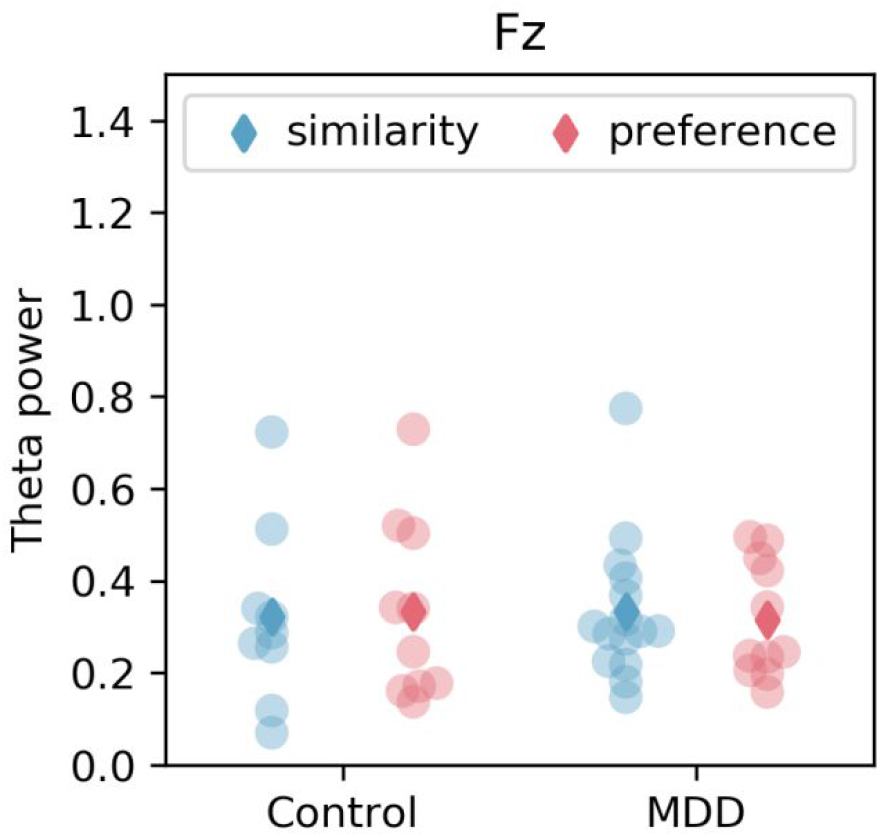
Theta band oscillatory power at the Fz electrode as a function of condition in MDD and control groups. Dots represent values from individuals in the similarity (blue) and preference (red) conditions. Group means are indicated by diamonds. No differences are observed.

**Supplementary figure 2:**
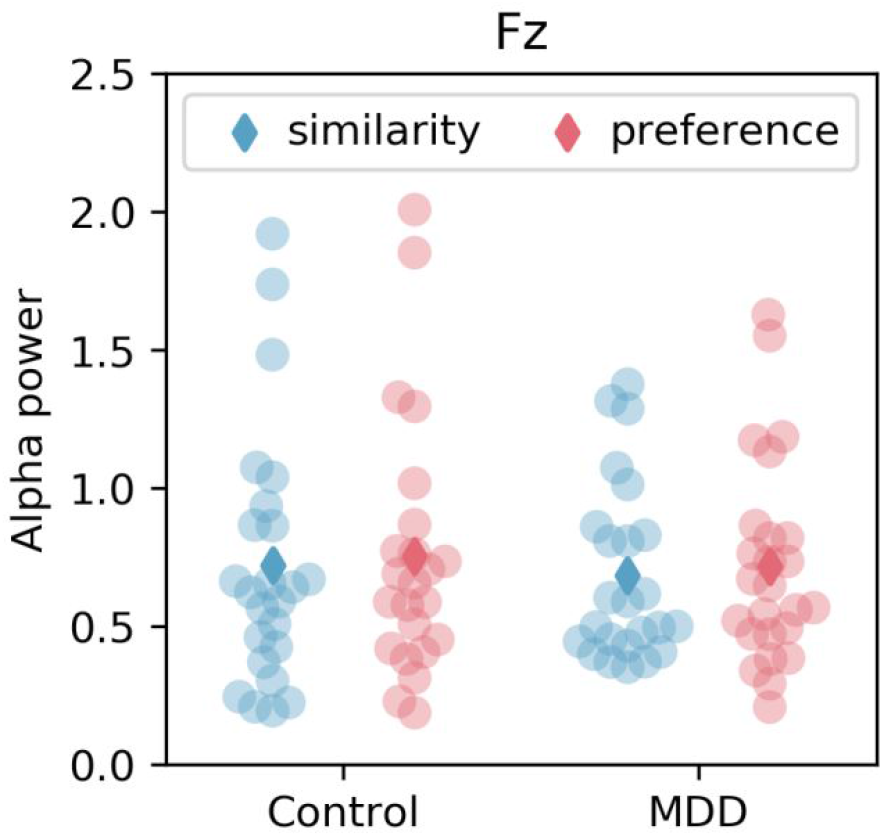
Alpha band oscillatory power at the Fz electrode as a function of condition in MDD and control groups. Dots represent values from individuals in the similarity (blue) and preference (red) conditions. Group means are indicated by diamonds. No differences are observed.

**Supplementary figure 3:**
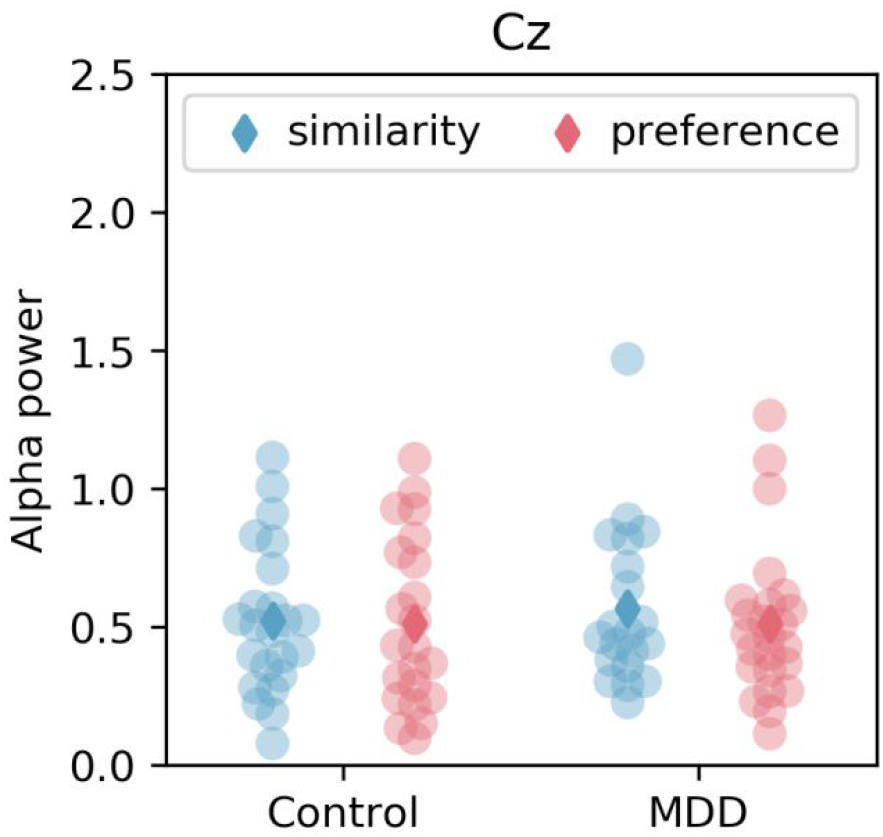
Alpha band oscillatory power at the Cz electrode as a function of condition in MDD and control groups. Dots represent values from individuals in the similarity (blue) and preference (red) conditions. Group means are indicated by diamonds. No differences are observed.

## Notes

### Competing Interest Statement

The authors have declared no competing interest.

### Author Declarations

Taipei Medical University JIRB

## References

Abela, J.R.Z., Hankin, B.L., 2011. Rumination as a vulnerability factor to depression during the transition from early to middle adolescence: a multiwave longitudinal study. J. Abnorm. Psychol. 120, 259–271. https://doi.org/10.1037/a0022796

Auerbach, R.P., Bondy, E., Stanton, C.H., Webb, C.A., Shankman, S.A., Pizzagalli, D.A., 2016. Self-referential processing in adolescents: Stability of behavioral and ERP markers. Psychophysiology 53, 1398–1406. https://doi.org/10.1111/psyp.12686

Auerbach, R.P., Stanton, C.H., Proudfit, G.H., Pizzagalli, D.A., 2015. Self-referential processing in depressed adolescents: A high-density event-related potential study. J. Abnorm. Psychol. 124, 233–245. https://doi.org/10.1037/abn0000023

Benau, E.M., Hill, K.E., Atchley, R.A., O’Hare, A.J., Gibson, L.J., Hajcak, G., Ilardi, S.S., Foti, D., 2019. Increased neural sensitivity to self-relevant stimuli in major depressive disorder. Psychophysiology 56, e13345. https://doi.org/10.1111/psyp.13345

Berman, M.G., Peltier, S., Nee, D.E., Kross, E., Deldin, P.J., Jonides, J., 2011. Depression, rumination and the default network. Soc. Cogn. Affect. Neurosci. 6, 548–555. https://doi.org/10.1093/scan/nsq080

Brainard, D.H., 1997. The Psychophysics Toolbox. Spat. Vis. 10, 433–436.

Delorme, A., Makeig, S., 2004. EEGLAB: an open source toolbox for analysis of single-trial EEG dynamics including independent component analysis. J. Neurosci. Methods 134, 9–21. https://doi.org/10.1016/j.jneumeth.2003.10.009

Disner, S.G., Beevers, C.G., Haigh, E.A.P., Beck, A.T., 2011. Neural mechanisms of the cognitive model of depression. Nat. Rev. Neurosci. 12, 467–477. https://doi.org/10.1038/nrn3027

Donoghue, T., Haller, M., Peterson, E.J., Varma, P., Sebastian, P., Gao, R., Noto, T., Lara, A.H., Wallis, J.D., Knight, R.T., Shestyuk, A., Voytek, B., 2020. Parameterizing neural power spectra into periodic and aperiodic components. Nat. Neurosci. 23, 1655–1665. https://doi.org/10.1038/s41593-020-00744-x

Ehinger, B., 2018. EEGVIS.

Fast, L.A., Funder, D.C., 2010. Gender differences in the correlates of self-referent word use: authority, entitlement, and depressive symptoms. J. Pers. 78, 313–338. https://doi.org/10.1111/j.1467-6494.2009.00617.x

Gotlib, I.H., Kasch, K.L., Traill, S., Joormann, J., Arnow, B.A., Johnson, S.L., 2004. Coherence and specificity of information-processing biases in depression and social phobia. J. Abnorm. Psychol. 113, 386–398. https://doi.org/10.1037/0021-843X.113.3.386

Grimm, S., Boesiger, P., Beck, J., Schuepbach, D., Bermpohl, F., Walter, M., Ernst, J., Hell, D., Boeker, H., Northoff, G., 2009. Altered negative BOLD responses in the default-mode network during emotion processing in depressed subjects. Neuropsychopharmacology 34, 932–843. https://doi.org/10.1038/npp.2008.81

Hamilton, J.P., Farmer, M., Fogelman, P., Gotlib, I.H., 2015. Depressive Rumination, the Default-Mode Network, and the Dark Matter of Clinical Neuroscience. Biol. Psychiatry, Depression 78, 224–230. https://doi.org/10.1016/j.biopsych.2015.02.020

Herbert, C., Pauli, P., Herbert, B.M., 2011. Self-reference modulates the processing of emotional stimuli in the absence of explicit self-referential appraisal instructions. Soc. Cogn. Affect. Neurosci. 6, 653–661. https://doi.org/10.1093/scan/nsq082

Holdaway, A.S., Luebbe, A.M., Becker, S.P., 2018. Rumination in relation to suicide risk, ideation, and attempts: Exacerbation by poor sleep quality? J. Affect. Disord. 236, 6–13. https://doi.org/10.1016/j.jad.2018.04.087

Huang, L.J., Wu, C.Y., Wu, C.H., Huang, P.S., Yeh, H.H., Yang, Y.H., Fang, Y.C., 2015. Validation of the Ruminative Response Scale-Chinese Version (RRS-C) for Persons with Depression in Taiwan. Taiwan. J Psychiatry 29, 119–131.

JASP Team, 2020. JASP.

Johnson, S.C., Schmitz, T.W., Kawahara-Baccus, T.N., Rowley, H.A., Alexander, A.L., Lee, J., Davidson, R.J., 2005. The cerebral response during subjective choice with and without self-reference. J. Cogn. Neurosci. 17, 1897–1906. https://doi.org/10.1162/089892905775008607

Kaiser, R.H., Whitfield-Gabrieli, S., Dillon, D.G., Goer, F., Beltzer, M., Minkel, J., Smoski, M., Dichter, G., Pizzagalli, D.A., 2016. Dynamic Resting-State Functional Connectivity in Major Depression. Neuropsychopharmacol. Off. Publ. Am. Coll. Neuropsychopharmacol. 41, 1822–1830. https://doi.org/10.1038/npp.2015.352

Knyazev, G., 2013. EEG Correlates of Self-Referential Processing. Front. Hum. Neurosci. 7. https://doi.org/10.3389/fnhum.2013.00264

Kuiper, N.A., Derry, P.A., 1982. Depressed and nondepressed content self-reference in mild depressives. J. Pers. 50, 67–80. https://doi.org/10.1111/j.1467-6494.1982.tb00746.x

Lemogne, C., Mayberg, H., Bergouignan, L., Volle, E., Delaveau, P., Lehéricy, S., Allilaire, J.-F., Fossati, P., 2010. Self-referential processing and the prefrontal cortex over the course of depression: a pilot study. J. Affect. Disord. 124, 196–201. https://doi.org/10.1016/j.jad.2009.11.003

Lopez-Calderon, J., Luck, S.J., 2014. ERPLAB: an open-source toolbox for the analysis of event-related potentials. Front. Hum. Neurosci. 8, 213. https://doi.org/10.3389/fnhum.2014.00213

Lu, M., 2002. Reliability and validity of the Chinese version of the Beck Depression Inventory-II. Taiwan. J Psychiatry 16, 301–310.

Mason, N.L., Kuypers, K.P.C., Müller, F., Reckweg, J., Tse, D.H.Y., Toennes, S.W., Hutten, N.R.P.W., Jansen, J.F.A., Stiers, P., Feilding, A., Ramaekers, J.G., 2020. Me, myself, bye: regional alterations in glutamate and the experience of ego dissolution with psilocybin. Neuropsychopharmacol. Off. Publ. Am. Coll. Neuropsychopharmacol. 45, 2003–2011. https://doi.org/10.1038/s41386-020-0718-8

Mennen, A.C., Norman, K.A., Turk-Browne, N.B., 2019. Attentional bias in depression: understanding mechanisms to improve training and treatment. Curr. Opin. Psychol. 29, 266–273. https://doi.org/10.1016/j.copsyc.2019.07.036

Mensen, A., Khatami, R., 2013. Advanced EEG analysis using threshold-free cluster-enhancement and non-parametric statistics. NeuroImage 67, 111–118. https://doi.org/10.1016/j.neuroimage.2012.10.027

Michalak, J., Hölz, A., Teismann, T., 2011. Rumination as a predictor of relapse in mindfulness-based cognitive therapy for depression. Psychol. Psychother. 84, 230–236. https://doi.org/10.1348/147608310X520166

Miskowiak, K.W., Larsen, J.E., Harmer, C.J., Siebner, H.R., Kessing, L.V., Macoveanu, J., Vinberg, M., 2018. Is negative self-referent bias an endophenotype for depression? An fMRI study of emotional self-referent words in twins at high vs. low risk of depression. J. Affect. Disord. 226, 267–273. https://doi.org/10.1016/j.jad.2017.10.013

Mu, Y., Han, S., 2013. Neural oscillations dissociate between self-related attentional orientation versus evaluation. NeuroImage 67, 247–256. https://doi.org/10.1016/j.neuroimage.2012.11.016

Nakao, T., Bai, Y., Nashiwa, H., Northoff, G., 2013a. Resting-state EEG power predicts conflict-related brain activity in internally guided but not in externally guided decision-making. NeuroImage 66, 9–21. https://doi.org/10.1016/j.neuroimage.2012.10.034

Nakao, T., Matsumoto, T., Morita, M., Shimizu, D., Yoshimura, S., Northoff, G., Morinobu, S., Okamoto, Y., Yamawaki, S., 2013b. The Degree of Early Life Stress Predicts Decreased Medial Prefrontal Activations and the Shift from Internally to Externally Guided Decision Making: An Exploratory NIRS Study during Resting State and Self-Oriented Task. Front. Hum. Neurosci. 7, 339. https://doi.org/10.3389/fnhum.2013.00339

Nejad, A.B., Fossati, P., Lemogne, C., 2013. Self-Referential Processing, Rumination, and Cortical Midline Structures in Major Depression. Front. Hum. Neurosci. 7. https://doi.org/10.3389/fnhum.2013.00666

Nolen-Hoeksema, S., Wisco, B.E., Lyubomirsky, S., 2008. Rethinking Rumination. Perspect. Psychol. Sci. J. Assoc. Psychol. Sci. 3, 400–424. https://doi.org/10.1111/j.1745-6924.2008.00088.x

Philippi, C.L., Cornejo, M.D., Frost, C.P., Walsh, E.C., Hoks, R.M., Birn, R., Abercrombie, H.C., 2018. Neural and behavioral correlates of negative self-focused thought associated with depression. Hum. Brain Mapp. 39, 2246–2257. https://doi.org/10.1002/hbm.24003

Qin, P., Northoff, G., 2011. How is our self related to midline regions and the default-mode network? NeuroImage 57, 1221–33. https://doi.org/10.1016/j.neuroimage.2011.05.028

Qin, P., Wang, M., Northoff, G., 2020. Linking bodily, environmental and mental states in the self—A three-level model based on a meta-analysis. Neurosci. Biobehav. Rev. 115, 77–95. https://doi.org/10.1016/j.neubiorev.2020.05.004

Schoofs, H., Hermans, D., Raes, F., 2010. Brooding and Reflection as Subtypes of Rumination: Evidence from Confirmatory Factor Analysis in Nonclinical Samples using the Dutch Ruminative Response Scale. J. Psychopathol. Behav. Assess. 32, 609–617. https://doi.org/10.1007/s10862-010-9182-9

Sheehan, D.V., Lecrubier, Y., Sheehan, K.H., Amorim, P., Janavs, J., Weiller, E., Hergueta, T., Baker, R., Dunbar, G.C., 1998. The Mini-International Neuropsychiatric Interview (M.I.N.I.): the development and validation of a structured diagnostic psychiatric interview for DSM-IV and ICD-10. J. Clin. Psychiatry 59 Suppl 20, 22-33;quiz 34-57.

Sheline, Y.I., Barch, D.M., Price, J.L., Rundle, M.M., Vaishnavi, S.N., Snyder, A.Z., Mintun, M.A., Wang, S., Coalson, R.S., Raichle, M.E., 2009. The default mode network and self-referential processes in depression. Proc. Natl. Acad. Sci. 106, 1942–1947. https://doi.org/10.1073/pnas.0812686106

Shestyuk, A.Y., Deldin, P.J., 2010. Automatic and strategic representation of the self in major depression: trait and state abnormalities. Am. J. Psychiatry 167, 536–544. https://doi.org/10.1176/appi.ajp.2009.06091444

Smith, J.M., Alloy, L.B., 2009. A roadmap to rumination: A review of the definition, assessment, and conceptualization of this multifaceted construct. Clin. Psychol. Rev. 29, 116–128. https://doi.org/10.1016/j.cpr.2008.10.003

Spasojevic, J., Alloy, L.B., 2001. Rumination as a common mechanism relating depressive risk factors to depression. Emot. Wash. DC 1, 25–37.

Spinhoven, P., van Hemert, A.M., Penninx, B.W., 2018. Repetitive negative thinking as a predictor of depression and anxiety: A longitudinal cohort study. J. Affect. Disord. 241, 216–225. https://doi.org/10.1016/j.jad.2018.08.037

Treynor, W., Gonzalez, R., Nolen-Hoeksema, S., 2003. Rumination Reconsidered: A Psychometric Analysis. Cogn. Ther. Res. 27, 247–259. https://doi.org/10.1023/A:1023910315561

Watkins, E.R., 2008. Constructive and Unconstructive Repetitive Thought. Psychol. Bull. 134, 163–206. https://doi.org/10.1037/0033-2909.134.2.163

Yoshimura, S., Okamoto, Y., Onoda, K., Matsunaga, M., Ueda, K., Suzuki, S., Shigetoyamawaki, null, 2010. Rostral anterior cingulate cortex activity mediates the relationship between the depressive symptoms and the medial prefrontal cortex activity. J. Affect. Disord. 122, 76–85. https://doi.org/10.1016/j.jad.2009.06.017

Young, K.D., Friedman, E.S., Collier, A., Berman, S.R., Feldmiller, J., Haggerty, A.E., Thase, M.E., Siegle, G.J., 2020. Response to SSRI intervention and amygdala activity during self-referential processing in major depressive disorder. NeuroImage Clin. 28, 102388. https://doi.org/10.1016/j.nicl.2020.102388

Zamoscik, V., Huffziger, S., Ebner-Priemer, U., Kuehner, C., Kirsch, P., 2014. Increased involvement of the parahippocampal gyri in a sad mood predicts future depressive symptoms. Soc. Cogn. Affect. Neurosci. 9, 2034–2040. https://doi.org/10.1093/scan/nsu006

Zhu, X., Wang, X., Xiao, J., Liao, J., Zhong, M., Wang, W., Yao, S., 2012. Evidence of a dissociation pattern in resting-state default mode network connectivity in first-episode, treatment-naive major depression patients. Biol. Psychiatry 71, 611–617. https://doi.org/10.1016/j.biopsych.2011.10.035

